# Antibiotic Resistance Trends Among Out-patients With Urinary Tract Infections In North Western Tanzania

**DOI:** 10.1101/2025.05.14.25327543

**Authors:** E. Magembe, S. Mapunjo, E. Mayenga, J. Shao, C. Lubega, K. Makhaola, I. Lumu, Tanzania Fleming Fund Fellowship consortium

## Abstract

**BACKGROUND:** Urinary tract infections (UTIs) are among the most common infections in the community and hospital settings and *Enterobacteriaceae*, are responsible for most infections. This study determined the prevalence and resistance trends of *E. coli* and *K. pneumoniae* to fluroquinolones and cephalosporins among out-patients diagnosed with urinary infections at a Zonal tertiary hospital in Tanzania

**METHODS:** This was a prospective cross sectional time series conducted in northern Tanzania and enrolled all out patients presenting with UTI symptoms. The study conducted for a period of six months between march 2021 to September 2021.

**RESULTS:** During the study period1582 patients were enrolled, the mean age was 20.2(SD 22.2) years and 883(55.8%) were female. The prevalence of *E. coli* was higher in female patients at 12.0% compared 6.7% in male. Both *E. coli* and *K. pneumoniae* were most prevalent in patients over >45 years at 13.3% and 3.2% respectively. *E*.*coli* resistance to Ceftriaxone, cefepime, and ciprofloxacin was 41.0%, 36.8% and 51.0% respectively. Resistance *K. pneumoniae* to ceftriaxone, cefepime, and ciprofloxacin was shown to be 52.8%, 47.4% and 28.2% respectively. Meanwhile, 76/192(39.6%) isolates were identified as ESBL.

**CONCLUSIONS:** In this setting empirical treatment of urinary tract infections with ciprofloxacin a commonly prescribed antibiotics for this diagnosis may not work in over half of patients when *E,coli* is the causative pathogen. Moreover, *E. coli*, resistance of 43% to ceftriaxone implies that patients who do not respond to initial therapy with ciprofloxacin, are at a risk of not responding to subsequent therapy necessitating the use of reserve antibiotics.

## INTRODUCTION

Antimicrobial resistance is an emerging global public health and economic problem (1,2,3), associated with significant mortality and morbidity (4,5). It is estimated that by 2050, global mortality from antimicrobial resistance will reach 10 million deaths annually (5,6), and cost the global economy over 100 trillion dollars each year (6). Worldwide, *E. coli* and *K. pneumoniae* are the major causes of community infections, such as urinary tract and intra-abdominal infections. Antibiotic-resistant strains of *Klebsiella pneumoniae*, are prevalent worldwide, posing substantial challenges as they are often implicated in severe and life-threatening infections (7). Urinary tract infections (UTIs) rank among the most frequently encountered bacterial infections in community and hospital settings (8) UTIs, particularly uncomplicated cystitis in women, are a frequent cause of antibiotic use in the community and in hospital setting (9).

Enterobacteriaceae, particularly E. coli, are the main pathogens, responsible for >80% of episodes of UTIs where a pathogen is identified and thus emphasizing their importance in antimicrobial surveillance (9,10). According to WHO, resistance *E. coli* isolates from urine samples to ceftriaxone ranged from 6% to 80%, while *K. pneumoniae* resistance to ceftriaxone ranged between 4 % and 90% in similar samples (11). In Tanzania, the AMR epidemiological landscape mirrors global trends, with studies indicating high resistance rates among *E. coli* and *K. pneumoniae* to commonly prescribed antibiotics, notably cephalosporins and fluoroquinolones (12). The Global Antimicrobial Resistance Partnership (GARP)-Tanzania Working Group AMR situation analysis reported that E. coli from urinary tract infections showed a 90% resistance to Ampicillin and 30-50% resistance to other common antibiotics. Extended-Spectrum Beta Lactamase (ESBL), which causes resistance to all beta lactam antibiotics, was found in 25-40% of *E. coli* (community and hospital sources) with more than 50% in children (13).

Fluoroquinolones, cephalosporins and β lactamase inhibitor combinations have been commonly used as first-line treatment options for these pathogens. Third-generation cephalosporins are now used widely, due to an increase in fluoroquinolone resistance in many countries (11,15). In addition, *Enterobacteriaceae* that produce extended-spectrum beta-lactamases (ESBL) have become endemic in many parts of the world, including Tanzania (10.) thereby limiting available treatment options and heightening the risk of therapeutic failure (16).The 2017 edition of Standard Treatment Guidelines (STG/NEMLIT) recommended the use of ciprofloxacin, amoxicillin +clavulanic acid and ceftriaxone in managing UTIs (14.) but there are concerns about the emergence and spread of resistance strains This study, therefore, was conducted to determine the prevalence and resistance trends of *E. coli* and *K. pneumoniae* to fluroquinolones and cephalosporins among out-patients diagnosed with urinary infections at a Zonal tertiary hospital in Tanzania.

## METHODOLOGY

This was a prospective cross-sectional time series study conducted for seven months between March and September 2021. The study included all outpatients diagnosed with Urinary tract infections (UTIs) at Bugando Medical Centre (BMC). BMC is a specialist and university teaching hospital for the Lake and Western zones of the United Republic of Tanzania. BMC is a tertiary referral hospital for eight regions and serves a catchment population of over 14 million people. The microbiology laboratory at BMC an ISO 15189 accredited laboratory and participates as a sentinel site for ARM surveillance.

### Sample collection and Laboratory procedures

Midstream urine samples were collected by the patients after instructions, following local procedures, and submitted to the laboratory within 2 hours after collection. Caretakers of children under the age of 5 years were given instructions to collect the urine sample of children and submit it to the testing laboratory within the same time. All samples were processed following local Standard operating procedures (SOPs) for urine culture, inoculating 10 microliters on Blood Agar and MacConkey Agar (Himedia, India) at BMC laboratory. In this study, urine samples with >10^5^ CFU/ml growth were considered positive (significant growth), a positive culture with one organism was considered a pure culture, and a growth of 2 pathogens, one or both with ≥10^5^ CFU/ml growth, was also considered as positive. Both isolates were subsequently sub-cultured separately to obtain pure culture. A negative urine culture was defined as any growth <10^5^ CFU/ml (no significant growth) and those without any bacterial growth. A urine sample showing more than two pathogens growth was considered contaminated, and the patient was instructed to provide another sample. Conventional biochemical identification method (Triple Sugar Iron (TSI), Sulfide Indole Motility (SIM), Citrate, Oxidase and Urease test) (Himedia, India) were done on appropriate quality controlled in-house prepared media by using established SOPs. Antibiotic susceptibility testing for ciprofloxacin 5μg, ceftriaxone 30μg, cefipime 30μg, ceftazidime 30μg, and amoxacillin/clavulanic acid 20/10μg, (Himedia, India) was performed to all identified isolates based on Clinical Laboratory Standard Institute guidelines (17) and 0.5 McFarland inoculum on Mueller Hinton agar (Himedia, India).

Phenotypic ESBL production was determined based on the disc approximation method where ceftriaxone 30μg, ceftazidime 30μg and Amoxicillin/Clavulanic acid 20/10μg placed 20mm apart on Mueller Hinton Agar to estimate ESBL prevalence in *E. coli* and *K. pneumoniae*. A zone difference of >5mm was considered as ESBL positive. ATCC bacterial strains *E. coli* ATCC 25922 (Susceptible wild type B-lactamase negative), and *K. pneumoniae* ATCC 700603 (ESBL producing strain, SHV-18) were used as control strains for media quality assurance, biochemical identification, AST, and ESBL testing. The *E. coli* and *K. pneumoniae* isolates identified from urine samples collected from outpatients at BMC in Mwanza city were transported in cold chain to NPHL in Dar es Salaam for further confirmation and storage.

### Data management and statistical analysis

We collected information on age, sex, specimen date, presenting symptoms, and a history of infection in the last three months. Clinical and demographic information, and microbiology data were entered into WHONET Software. Frequencies, percentages, and standard deviations were used to summarize the clinical characteristics of outpatients presenting with UTI. The proportion for *K. pneumoniae* and *E. coli* isolates recovered from urine samples, and along with the antibiotic susceptibility profile, was calculated. In some cases we combine intermediate and resistant results into a single group and report as non-susceptibility. The resistance percentage and 95% confidence interval of *K. pneumoniae and E. coli against Fluoroquinolones and* cephalosporins derived. The data presented is presented according to the CLSI guides(18) and analysis was done using WHONET and R studio software.

### Ethical Consideration

Ethical clearance for the study was issued by the National Institute for Medical Research-Reference code (NIMR/HQ/R.8a/Vol.IX/3554.

## RESULTS

During the study period (March to September 2021), a total of 1582 patients were enrolled and 1582 non-repetitive urine samples were analysed. The mean age was 20.2 years (SD 22.2). Most samples 883(55.8%) were from Female patients (See Table 1). Out of 1582 processed specimens during the study period, 400 (25.2%) samples were culture positive and 1182 (74.7%) were culture negative or showed no significant growth. *E. coli* was isolated from 153 (9.7%) and *K. pneumoniae* from 39 (2.5%) while 208(13.1%) were identified as others isolates (supplementary table 1). The prevalence of *E. coli* was higher in female patients at 12.0% compared 6.7% in male. Meanwhile, the prevalence of *K. pneumoniae* was similar in both sexes. Both *E. coli* and *K. pneumoniae were* the most prevalent in patients aged over 45 years at 13.3% and 3.2% respectively (supplementary table 1)

**Table 1.** Demographic and Clinical characteristics of outpatients Patients presenting at BMC with suspected UTI, March - April 2022.

| Characteristic | N (%) |
| --- | --- |
| **Age** | 20.2(sd-22.2) |
| 0-14 | 957 (60.5%) |
| 15-45 | 340 (21.5%) |
| >45 | 285 (18.0%) |
| **Sex** |  |
| Female | 883 (55.8%) |
| Male | 699 (44.2%) |
| **Symptoms** |  |
| Abdominal pain |  |
| No | 999 (63.1%) |
| Yes | 579 (36.6%) |
| Missing | 4 (0.3%) |
| **Dysuria** |  |
| No | 1542 (97.5%) |
| Yes | 36 (2.3%) |
| Missing | 4 (0.3%) |
| **Antibiotic History** |  |
| No | 1441(91.1%) |
| Yes | 134(8.5%) |
| Missing | 7(0.4%) |
| **Recurrent UTI** |  |
| No | 1488(94%) |
| Yes | 90(5.7%) |
| Missing | 4(0.3%) |

### Susceptibility patterns and trends

Of 153 *E. coli* isolates, 151 were tested for susceptibility to third-generation cephalosporins (ceftriaxone), 152 to fourth-generation cephalosporins (cefepime), and 153 were tested for fluoroquinolones (ciprofloxacin). Resistance to Ceftriaxone was 41.0%, cefepime resistance was observed in 36.8% and for ciprofloxacin, 51.0% resistance (Table 2). Of the 39 *K. pneumoniae* isolates 36 were tested for third-cephalosporins (ceftriaxone), 38 isolates tested for fourth-generation cephalosporins (cefepime), and all 39 isolates were tested against fluoroquinolones (ciprofloxacin), for which resistance was shown to be 52.8%, 47.4% and 28.2% respectively.

**Table 2.**
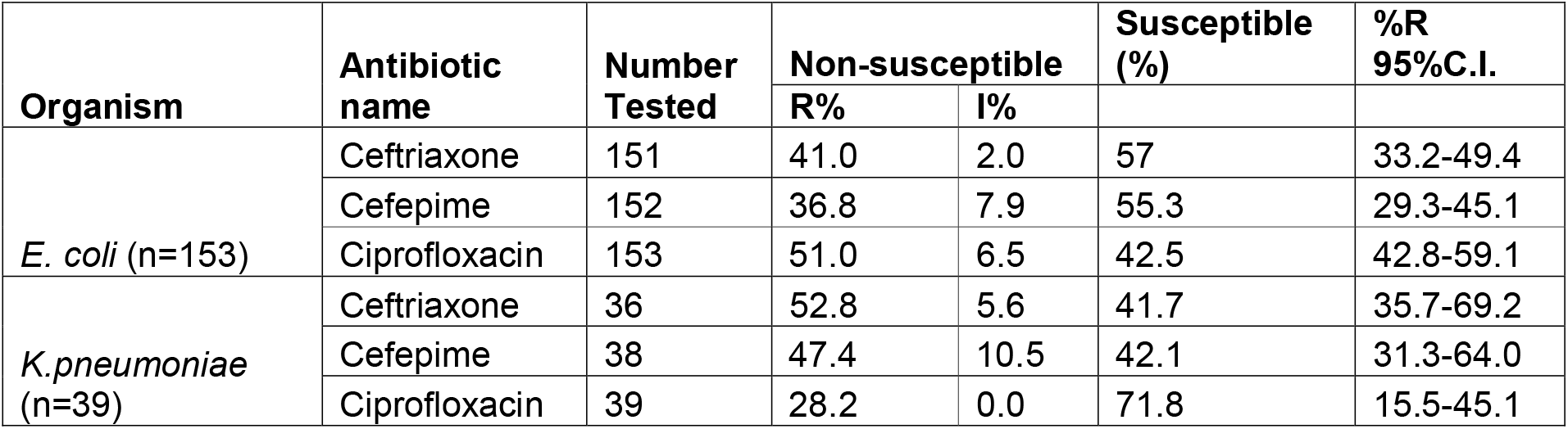
Resistance profiles E. coli and K. pneumoniae for ceftriaxone, cefepime and ciprofloxacin, BMC, March – September 2021.

Higher resistance was observed in both *K. pneumoniae* isolates from female patients compared to isolates from male patients (Figure 1) Up to 36.2% of *E*.*coli* were resistant to Cefepime and 52.8% to Ciprofloxacin while for *K. pneumoniae* 54.5% resistance to cefepime(Table1). Meanwhile, 76/192(39.6%) isolates were ESBL phenotypes of which 57/192 (29.6%) were *E. coli* and 19/192 (9.9%) were *K. pneumoniae* isolated. Figure 2 shows the resistance trend of *E*.*coli* to ciprofloxacin, ceftriaxone and cefepime over 7 months. Generally, the resistance of *E. coli* to ciprofloxacin remained higher compared to ceftriaxone and cefepime the entire period.

**Figure 1.**
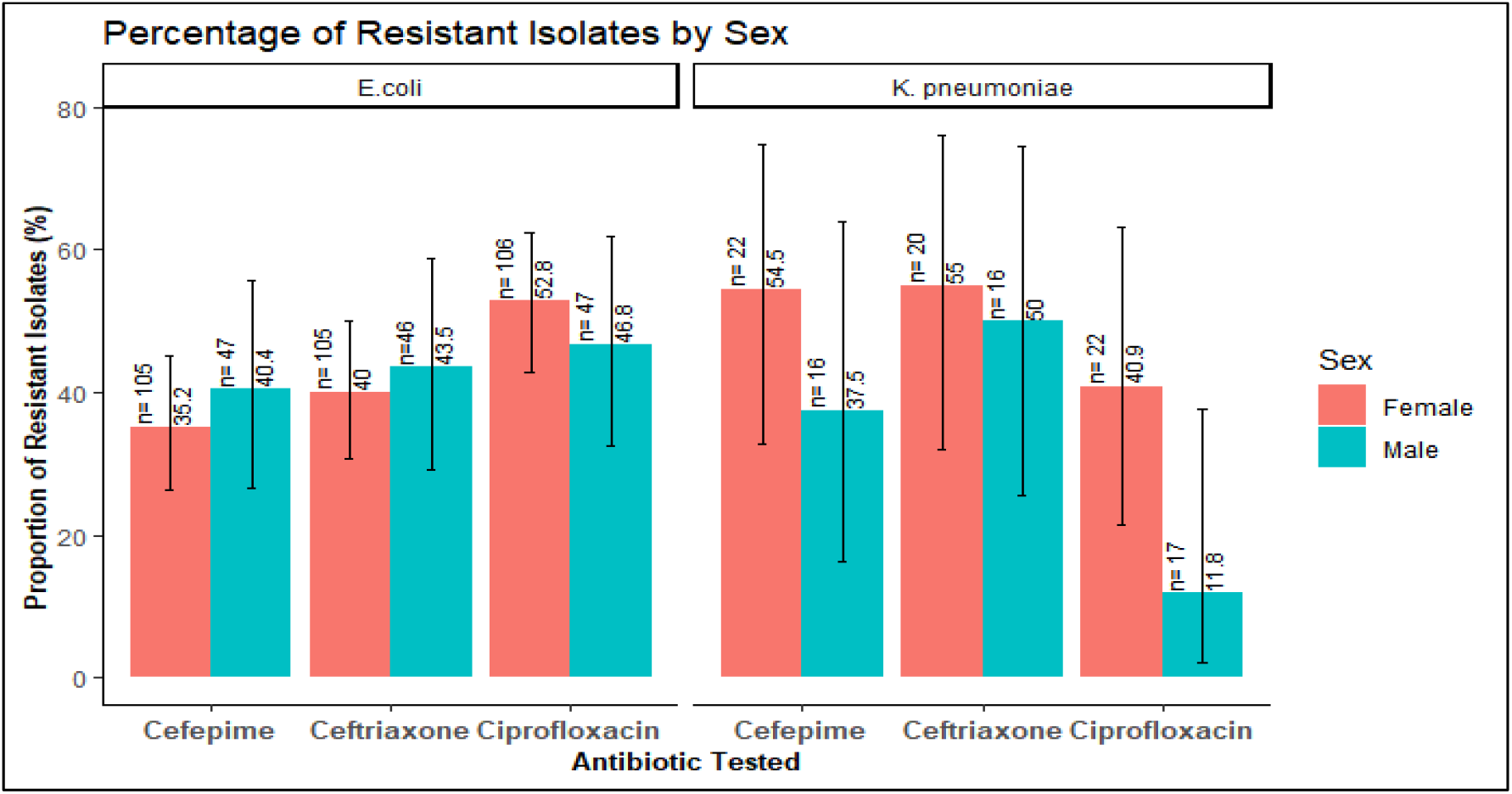
Resistance of E. coli and K. pneumoniae by sex, BMC, March – September 2021.

**Figure 2.**
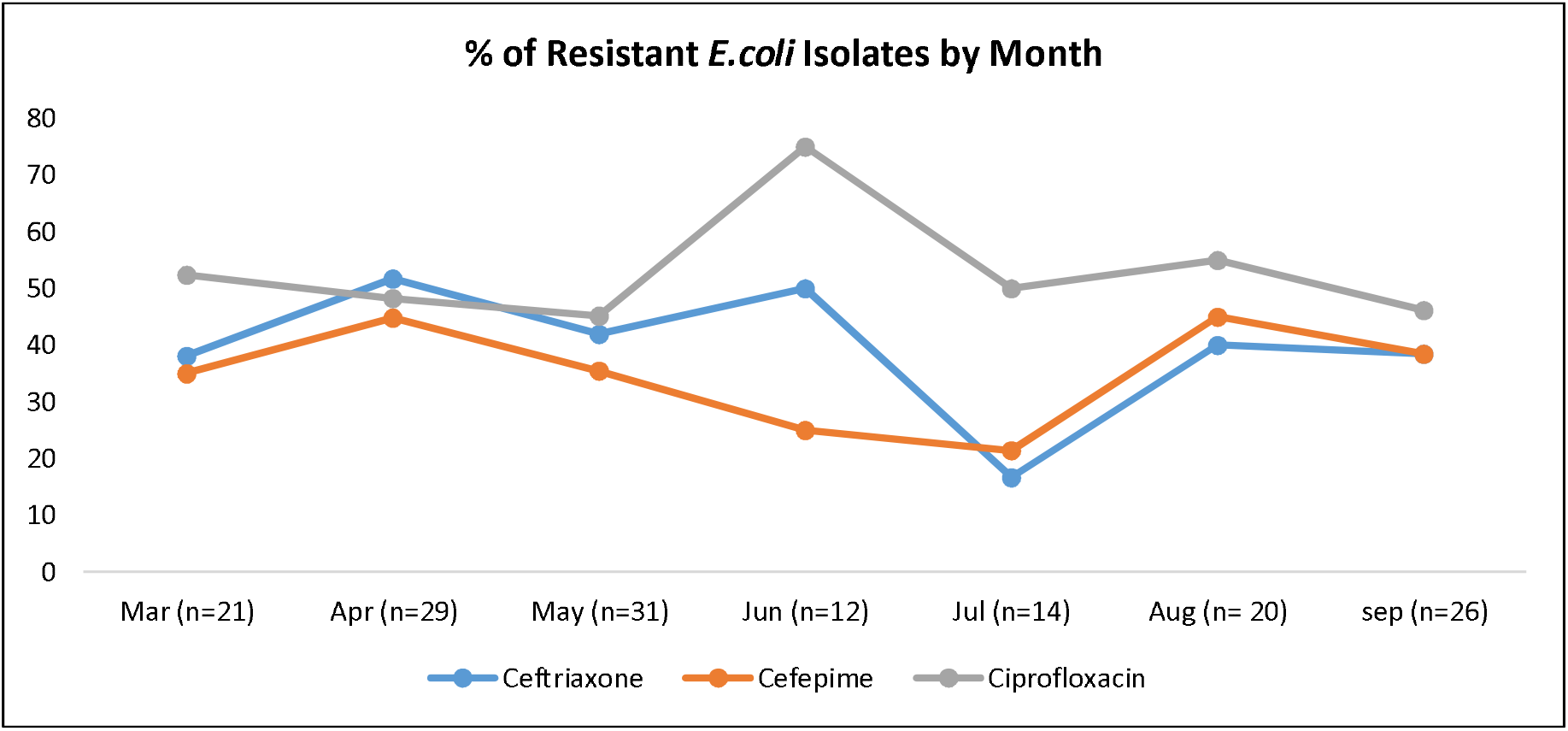
Resistance trends of E. coli isolates by month, BMC, March – September 2021.

## DISCUSSION

This study was conducted to investigate the prevalence and trends of resistance in uropathogens isolated from patients presenting with UTIs, with the ultimate goal of extrapolating this data to guide policymaking. Moreover, the investigators hoped to, highlight the need and demonstrate the feasibility of, a routine surveillance program at a sentinel in Tanzania. This study has shown that among UTI cases, at a tertiary referral hospital, *E. coli* was the most common causative pathogen in 9.7% (153) of cases, making it a representative uropathogen in AMR surveillance.

We observed a high incidence of *E*.*coli* associated infection in female patients 12004/100,000 samples which was higher compared to male patients 6723/100,000. This finding is in agreement with other studies explaining that UTI is more prevalent in female patients than in male patients (19). Also, the prevalence for *E. coli* and *K*.*pneumonie* (13.3% vs 3.2%) was higher in patients between patients >45 years compared to other age groups. This implies that of the 100000 patients aged >45 years that attend BMC with signs of UTI, this will translate to patients will 13333/samples and 3157/ be diagnosed with UTI due to *E. coli* and *K*.*pneumoniae* respectively. This observation would be due to anatomical differences in females that predispose women to UTI and also age-related change as UTI become more common with advancing age in both sexes (20). Additionally, we observed a high incidence of ESBL (48718/100000 isolate) in *K*.*pneumoniae* isolate compared to *E. coli* isolate(37253/100000 isolates). The high proportion of ESBL would be due to recurrent and frequent UTIs, especially in females and older patients necessitating frequent use of antibiotics.

*E. coli* and *K. pneumoniae* were the most common pathogen found in urine highlighting the need to continuously monitor their prevalence. These findings show high resistance rates to one of the second-line treatment options, according to the National Treatment Guidelines, and commonly used antibiotic for UTI treatment. A resistance rate of 51% for ciprofloxacin was shown among *E. coli* isolates, while this was 28.2% for *K*.*pneumoniae* isolates among patients treated for UTI. These findings imply that one in two of *E. coli* associated UTIs may not be responsive to empirical treatment of ciprofloxacin, while almost one-third of *K. pneumoniae* associated-UTIs may not be treated with Ciprofloxacin. The high resistance observed may be due to overuse of ciprofloxacin which until recently was the first line treatment for UTI according to National Guidelines (14). Additionally, urinary tract infections are more common in females than in males (19,20), leading to frequent use of antibiotics in this population group and consequently resulting in the high resistance rates of K. pneumonia observed in this study.

We observed a generally high resistance to ciprofloxacin throughout the study period followed by ceftriaxone this would be due to the increased use of these antibiotics in routine care. Additionally, Ciprofloxacin is easily accessible in the community and this could be a driving factor for its misuse, and subsequently driving factor to AMR (21). The rise of ciprofloxacin resistance in June is difficult to explain but the observation period was equally short to shade sufficient light on this. Although cefepime is not commonly used in routine care the resistance trends are concerning and may be a result of cross resistance to other generations of cephalosporins which are more readily available.

The study provided a sufficient number of isolates to highlight the AMR situation in Northern Tanzania among patients diagnosed or investigated for UTI. Moreover, this study was a prospective cross-sectional time series of ARM trends over 7 months. To our knowledge, this was the first time series conducted during the peak of the COVID-19 pandemic to investigate the resistance trends of ceftriaxone and ciprofloxacin in an outpatient settings. The current study highlights the significance of culture-guided therapy to improve patient care and antibiotic stewardship in an attempt to curb AMR in low and middle-income countries. On the other hand, the results this study should be interpreted with caution since the study was performed at a single tertiary referral hospital and there may be variations across the region. Additionally, the Zonal hospital receives patients with complicated medical histories, or that have been previously treated with antibiotics, thus overestimating the level of resistance. Lastly, no information on treatment outcomes was included in the current study. Of interest for future exploration could be a program investigating patient treatment outcome, as a measure to monitor effectiveness of empirical antibiotic treatment, and its effect on AMR in the indicator pathogens *E. coli* and *K. pneumoniae*.

In summary, in this setting empirical treatment of urinary tract infections with ciprofloxacin a commonly prescribed antibiotic for this diagnosis may not work in over half of patients where *E*.*coli* is isolated. Moreover, the *E. coli*, non-susceptibility rate of 43% to ceftriaxone -the second-line option, means that patients not responding to initial therapy with ciprofloxacin, may not respond to subsequent therapy necessitating the use of reserve antibiotics.

## Data Availability

The data is available via reasonable requests directed to the Tanzania Fleming Fund Fellowship consortium.

## Acknowledgement

ME, LI, conceptualised and designed the study and drafted the first manuscript.

LC LI, ME., Analysed and interpreted the data.

MS, SJ, LI, ME Wrote sections of the manuscript

LI, MK: supervised the writing of the manuscript and critically reviewed the manuscript We thank the study staff at Bugando Medical centre, Igembe Zechariah, and Peter Peter. Conflict of Interest: No conflict of interest to declare

## Tanzania Fleming Fund consortium

Nyambura Moremi., Kolader Marion., Schultsz Constance., Shumba.Edwin., Ondoa Pascale., Richard Walemwa., Beverly Egyir., Neema Kamala., Kayuni Gibbonce., Oskam Linda.

## References

1. WHO (2015) Global antimicrobial resistance surveillance system (GLASS): manual for early implementation. Geneva: WHO

2. Jonas OB, Irwin A, Berthe FC, Le Gall FG, Marquez PV. Drug-resistant infections: a threat to our economic future. World Bank Rep. 2017;2:1–32.

3. Founou RC, Founou LL, Essack SY. Clinical and economic impact of antibiotic resistance in developing countries: A systematic review and meta-analysis. PloS one. 2017 Dec 21;12(12):e0189621.

4. Murray CJ, Ikuta KS, Sharara F, Swetschinski L, Aguilar GR, Gray A, Han C, Bisignano C, Rao P, Wool E, Johnson SC. Global burden of bacterial antimicrobial resistance in 2019: a systematic analysis. The lancet. 2022 Feb 12;399(10325):629–55.

5. Naghavi M, Vollset SE, Ikuta KS, Swetschinski LR, Gray AP, Wool EE, Aguilar GR, Mestrovic T, Smith G, Han C, Hsu RL. Global burden of bacterial antimicrobial resistance 1990–2021: a systematic analysis with forecasts to 2050. The Lancet. 2024 Sep 28;404(10459):1199–226.

6. O’Neil, J. (2017) Review of Antimicrobial resistance: final report and recommendations. Available at: https://amr-review.org/sites/default/files/160525_Final%20paper_with%20cover.pdf (Accessed 01/05/2020)

7. Ndlovu T, Kgosietsile L, Motshwarakgole P, Ndlovu SI. Evaluation of potential factors influencing the dissemination of multidrug-resistant Klebsiella pneumoniae and alternative treatment strategies. Tropical Medicine and Infectious Disease. 2023 Jul 26;8(8):381.

8. Fasugba O, Gardner A, Mitchell BG, Mnatzaganian G. Ciprofloxacin resistance in community-and hospital-acquired Escherichia coli urinary tract infections: a systematic review and meta-analysis of observational studies. BMC infectious diseases. 2015 Dec;15:1–6.

9. Al-Zahrani J, Al Dossari K, Gabr AH, Ahmed AF, Al Shahrani SA, Al-Ghamdi S. Antimicrobial resistance patterns of Uropathogens isolated from adult women with acute uncomplicated cystitis. BMC microbiology. 2019 Dec;19:1–5.

10. Zanichelli, V. et al. (2019) ‘Antimicrobial resistance trends in Escherichia coli, Klebsiella pneumoniae and Proteus mirabilis urinary isolates from Switzerland: retrospective analysis of data from a national surveillance network over an 8-year period (2009-2016)’, Swiss medical weekly. NLM (Medline), 149, p. w20110. doi: 10.4414/smw.2019.20110.

11. WHO (2022) Global antimicrobial resistance and use surveillance system (GLASS) report 2022

12. Letara N, Ngocho JS, Karami N, Msuya SE, Nyombi B, Kassam NA, Skovbjerg S, Åhren C, Philemon R, Mmbaga BT. Prevalence and patient related factors associated with Extended-Spectrum Beta-Lactamase producing Escherichia coli and Klebsiella pneumoniae carriage and infection among pediatric patients in Tanzania. Scientific Reports. 2021 Nov 23;11(1):22759.

13. Ministry of Health (2017) The United Republic of Tanzania; National Action Plan on Antimicrobial Resistance 2017-2022’, United Republic of Tanzania

14. Ministry of Health Community Development Gender Elderly and Children (2017) ‘Standard Treatment Guidelines & National Essential Medicines List, Tanzania Mainland G’, p. 333 & 334.

15. White RT. Escherichia coli: placing resistance to third-generation cephalosporins and fluoroquinolones in Australia and New Zealand into perspective. Microbiology Australia. 2021 Sep 8;42(3):104–10.

16. Lee, S. et al. (2014) ‘Third-generation cephalosporin resistance of community-onset Escherichia coli and Klebsiella pneumoniae bacteremia in a secondary hospital’, Korean Journal of Internal Medicine, 29(1), pp. 49–56. doi: 10.3904/kjim.2014.29.1.49.

17. CLSI (2011) Performance standards for antimicrobial susceptibility testing; twenty first information supplement. vol. CLSI document M100-S21. Wayne: Clinical and Laboratory Standards Institute

18. Hindler, J.F. and Stelling, J. (2007) Analysis and presentation of cumulative antibiograms: a new consensus guideline from the Clinical and Laboratory Standards Institute. Clinical infectious diseases, 44(6), pp.867–873.

19. Odoki M, Almustapha Aliero A, Tibyangye J, Nyabayo Maniga J, Wampande E, Drago Kato C, Agwu E, Bazira J. Prevalence of bacterial urinary tract infections and associated factors among patients attending hospitals in Bushenyi district, Uganda. International journal of microbiology. 2019;2019(1):4246780.

20. Magliano E, Grazioli V, Deflorio L, Leuci AI, Mattina R, Romano P, Cocuzza CE. Gender and age-dependent etiology of community-acquired urinary tract infections. The scientific world journal. 2012;2012(1):349597.

21. Tchesnokova V, Larson L, Basova I, Sledneva Y, Choudhury D, Solyanik T, Heng J, Bonilla TC, Pham S, Schartz EM, Madziwa LT. Increase in the community circulation of ciprofloxacin-resistant Escherichia coli despite reduction in antibiotic prescriptions. Communications Medicine. 2023 Aug 12;3(1):110.

