## supplementary material for "Antibiotic Resistance Trends Among Out-patients With Urinary Tract Infections In North Western Tanzania"

***Supplementary Figure 1:*** *Prevalence of E. coli and K. pneumoniae by month among patients with suspected UTI at BMC, March – September 2021*

Culture Results by Month

|  | K. pneumoniae | E.coli | Other Pathogens | No growth or no significant growth | Samples Cultured |
| --- | --- | --- | --- | --- | --- |
| 2021-03-01 | 4 | 21 | 26 | 77 | 128 |
| 2021-04-01 | 6 | 29 | 38 | 219 | 292 |
| 2021-05-01 | 8 | 31 | 49 | 214 | 302 |
| 2021-06-01 | 6 | 12 | 30 | 172 | 220 |
| 2021-07-01 | 4 | 14 | 13 | 119 | 150 |
| 2021-08-01 | 3 | 20 | 24 | 159 | 206 |
| 2021-09-01 | 8 | 26 | 28 | 222 | 284 |

***Supplementary Table 1:*** *Prevalence of E. coli and K. pneumoniae among patients investigated UTI at BMC, March – September 2021*

|  | **Female**  **N=883(Prevalence)** | **Male**  **N=699(Prevalence)** | **Overall**  **N=1582(Prevalence)** |
| --- | --- | --- | --- |
| **Culture Results** |  |  |  |
| *E. coli* | 106 (12.0%) | 47(6.7%) | 153 (9.7%) |
| *K. pneumoniae* | 22 (2.5%) | 17(2.4%) | 39(2.5%) |
| Other Pathogens | 116 (13.1%) | 92(13.2%) | 208(13.1%) |

***Supplementary Table 2:*** *Prevalence of E. coli and K. pneumoniae by age group among patients with suspected UTI at BMC, March – September 2021*

|  | 0-14  N=957(Prevalence) | 15-45  N=340(Prevalence) | Above 45  N=285(Prevalence) | Overall  N=1582(Prevalence) |
| --- | --- | --- | --- | --- |
| Culture Results | | | | |
| *E. coli* | 85(8.9%) | 30(8.8%) | 38(13.3%) | 153 (9.7%) |
| *K. pneumoniae* | 27 (2.8%) | 3(0.9%) | 9 (3.2%) | 39 (2.5%) |
| Other Pathogens | 124(3.0%) | 35 (10.3%) | 49(17.2%) | 208(3.1%) |
